# Decoding the metabolic blockade effect: PFAS inhibition of organic anion transporters impairs VOC clearance and amplifies neurocognitive decline

**DOI:** 10.64898/2026.02.12.26346123

**Authors:** Lili Liang, Spencer Xinyi Zhang, Jenny J Lin

**Affiliations:** Division of Gastroenterology, Icahn School of Medicine at Mount Sinai, New York, NY, USA; Department of Psychiatry, Icahn School of Medicine at Mount Sinai, New York, NY, USA; Division of General Internal Medicine, Icahn School of Medicine at Mount Sinai, New York, NY, USA

**Author notes:** **Corresponding author:** Lili Liang.

**Keywords:** PFAS, Volatile Organic Compounds, Metabolic Blockade, Double Machine Learning, Molecular Docking, Synergistic Neurotoxicity

## Abstract

The co-occurrence of per- and polyfluoroalkyl substances (PFAS) and volatile organic compounds (VOCs) in industrial environments poses complex toxicological risks that standard additive models fail to capture. This study elucidates a novel “metabolic blockade” mechanism wherein PFAS competitively inhibits the renal excretion of VOC metabolites, thereby amplifying neurotoxic burdens. Utilizing a Double Machine Learning (DML) framework on data from National Health and Nutrition Examination Survey (2005–2020), we analyzed a final intersectional cohort of 1,975 participants. We identified a robust inhibition of VOC metabolite clearance by serum PFAS. Specifically, PFNA significantly suppressed the excretion of the benzene metabolite URXPMA (Causal *β*_*TMLE*_ = −0.219, *p* < 0.001), with efficacy dependent on perfluorinated chain length. Molecular docking simulations revealed the biophysical basis of this antagonism: long-chain PFNA exhibited superior binding affinity to the Organic Anion Transporter 1 (OAT1) (Δ*G* = −6.333 kcal/mol) compared to native VOC metabolites (Δ*G* = −4.957 kcal/mol), confirming high-affinity competitive inhibition at the renal interface. In a neurocognitive sub-cohort (N = 1,200), this interference translated into functional synergism; high-PFNA exposure magnified VOC-associated cognitive impairment by 1.5-fold and significantly exacerbated the negative association between VOC burden and processing speed (*β*_*int*_ = −0.263, *p* = 0.004). These findings define PFAS as a “metabolic amplifier” of co-contaminant toxicity, necessitating a paradigm shift toward mixture-based hazardous material regulations that account for transporter-level interactions.

## 1. Introduction

Per- and polyfluoroalkyl substances (PFAS) represent a class of synthetic fluorinated hydrocarbons characterized by exceptional thermal and chemical stability, posing a formidable challenge to global environmental management (Güzel, 2025; Nayak and Yamijala, 2024). Since the early 2000s, large-scale observational studies have confirmed the ubiquitous presence of PFAS in the hydrosphere and their significant potential for bioaccumulation in human circulation (Sunderland et al., 2019; De Silva et al., 2021). Long-chain congeners, such as perfluorooctane sulfonate (PFOS) and perfluorooctanoic acid (PFOA), possess high affinities for serum proteins and have been causally linked to a spectrum of adverse health outcomes, including immune dysregulation (DeWitt et al., 2012; Corsini et al., 2014), endocrine disruption (Di Nisio et al., 2022), and metabolic disruption (Niu et al., 2023). Despite increased global regulations on legacy PFAS, emerging alternatives and residual long-chain congeners continue to threaten human health through the persistent contamination of drinking water and food chains (Cousins et al., 2020; Figuière et al., 2025).

Concurrently, volatile organic compounds (VOCs) have emerged as primary pollutants of concern in industrialized urban zones. Recent spatial-environmental assessments indicate that in areas surrounding petrochemical complexes, hydrocarbon pollutants form a sustained pollution load, accumulating in soil-plant systems through atmospheric deposition and subsequently entering the human exposure pathway (Kazemi et al., 2026). The neurotoxicity of these compounds is extensively documented; specifically, chronic exposure to benzene and toluene is associated with impaired executive function and processing speed (Khan et al., 2021; Killin et al., 2016). Mechanistically, recent clinical evidence suggests that the intracellular accumulation of VOC metabolites induces oxidative stress and mitochondrial dysfunction within the brain microenvironment, acting as a key driver of neurodegeneration (Laganà et al., 2026; Hajjar et al., 2018; Molot et al., 2021). Furthermore, the interaction of these pollutants with microplastic vectors may further complicate their bioavailability and toxicological profiles in complex mixtures (Sofield et al., 2026).

However, current environmental risk assessment frameworks predominantly adhere to traditional mixture paradigms, such as toxic equivalency factors or additive models, which often overlook non-additive interactions arising from toxicokinetic interference (Braun and Gray, 2017; Taylor et al., 2016). This gap is critical given that combined exposures are an enduring priority for environmental health research (Carlin and Rider, 2024). We posit a novel “Metabolic Blockade” mechanism: PFAS, due to their structural similarity to fatty acids, may competitively bind to critical clearance portals, specifically Organic Anion Transporters (OATs) and Human Serum Albumin (HSA), inhibiting the renal elimination of VOC metabolites (Pelis and Wright, 2011; Louisse et al., 2024). This blockade could abnormally prolong the biological half-lives of non-persistent neurotoxicants, effectively lowering the physiological threshold for neurological damage in vulnerable populations (Constantinescu et al., 2025; Iyevhobu et al., 2025). Although PFAS-transporter interactions have been documented (Ryu et al., 2024), whether this molecular competition translates into impaired VOC clearance and synergistic neurotoxicity remains a critical knowledge gap.

To address this challenge, this study utilizes data from the National Health and Nutrition Examination Survey (NHANES, 2005–2020) to systematically evaluate the impact of PFAS on VOC metabolic clearance. We employ a Double Machine Learning (DML) framework to control for high-dimensional confounding, allowing for a robust estimation of the association between PFAS burden and VOC metabolite excretion (Chernozhukov et al., 2018). This epidemiological evidence is complemented by molecular docking simulations to verify the biophysical mechanism of competitive binding at OAT1 and HSA sites (Forli et al., 2016; Cheng and Ng, 2018). Finally, to assess the clinical downstream effects of this blockade, we constructed a targeted neurocognitive sub-cohort of older adults. Within this vulnerable population, we evaluate whether PFAS-induced metabolic interference translates into functional synergism, specifically amplifying declines in processing speed and memory. By integrating large-scale population data with computational toxicology, this multidisciplinary approach seeks to establish a new paradigm for investigating mechanistic interactions within complex chemical mixtures.

## 2. Materials and Methods

### 2.1 Study Population and Data Integration

As illustrated in the study flow diagram (Figure S1), the analytic sample was constructed through a rigorous sequential exclusion process initiated on the general adult population (aged ≥ 20 years). To ensure valid concurrent exposure assessment, we restricted the analysis to the intersection of participants possessing valid measurements for both serum PFAS and urinary VOC metabolites, a necessary constraint due to the independent subsampling schemes employed by NHANES. Furthermore, to mitigate physiological confounding arising from hydration status, participants with urinary creatinine concentrations less than 30 mg/dL or greater than 350 mg/dL were excluded to prevent bias from overly dilute or concentrated urine samples. The final analysis was subsequently stratified into two distinct datasets: a Tier 1 (Metabolic Cohort) comprising the general adult population (*N* = 1,975) utilized for the causal modeling of PFAS-VOC metabolic interactions, and a Tier 2 (Neurocognitive Sub-cohort) consisting of a targeted sample of older adults (aged ≥ 60 years) derived from the 2011–2014 cycles. To address missingness in covariates within the Tier 2 cohort, Multiple Imputation by Chained Equations (MICE) was employed with 10 iterations, yielding a finalized analytical sample of 1,200 participants.

### 2.2. Laboratory Measurements and Cognitive Assessment

Serum concentrations of PFOS, PFOA, perfluorononanoic acid (PFNA), and perfluorohexane sulfonate (PFHxS) were quantified using high-performance liquid chromatography-tandem mass spectrometry (HPLC-MS/MS). Urinary VOC metabolites serving as biomarkers for volatile organic compound exposure were quantified via ultra-high-performance liquid chromatography-tandem mass spectrometry (UPLC-MS/MS). Specifically, phenylmercapturic acid (PMA), N-acetyl-S-(benzyl)-L-cysteine (BMA), and 2-methylhippuric acid (2MHA) were measured as biomarkers for benzene, toluene, and xylene, respectively. All urinary concentrations were creatinine-adjusted to correct for hydration status. In the Tier 2 cohort, cognitive function was assessed using three standardized instruments: the Consortium to Establish a Registry for Alzheimer’s Disease (CERAD) Word Learning (immediate and delayed recall), the Animal Fluency test (verbal fluency), and the Digit Symbol Substitution Test (DSST) (processing speed). Raw scores were standardized to Z-scores to facilitate cross-domain comparisons.

### 2.3. Statistical Analysis

Metabolic associations between serum PFAS (treatment) and urinary VOC metabolites (outcome) in the Tier 1 cohort were estimated using a Double Machine Learning (DML) framework (Chernozhukov et al., 2018). Nuisance parameters were modeled using Random Forest regressors and Gradient Boosting machines to orthogonalize residuals. A 5-fold cross-fitting procedure was implemented to prevent overfitting. Additionally, Target Maximum Likelihood Estimation (TMLE) was applied to estimate the Average Treatment Effect (ATE) of PFAS quartiles on metabolite clearance. For the neurocognitive interaction analysis in Tier 2, multivariable linear regression models were fitted to test the “Metabolic Blockade” hypothesis. The model was specified as:

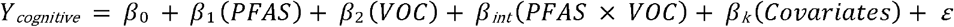

Significant negative interaction terms (*β*_*int*_) indicated synergistic toxicity. All models were adjusted for age, educational attainment, body mass index (BMI), smoking status, alcohol consumption, creatinine, and serum albumin. All analyses above were conducted using Python v3.12.12 and incorporated NHANES complex survey weights to account for the stratified multistage sampling design.

### 2.4. Molecular Docking Simulation

Molecular docking was performed using AutoDock Vina v1.2.5. The crystal structures of Human Serum Albumin (HSA, PDB ID: 1AO6) and Organic Anion Transporter 1 (OAT1, PDB ID: 6Y7W) were retrieved from the RCSB Protein Data Bank. Protein preparation involved the removal of crystallographic water molecules, addition of polar hydrogens, and assignment of Gasteiger charges. The search space was defined by a 20 × 20 × 20 Å grid box centered on the active binding sites (OAT1 center coordinates: *X* = −5.88, *Y* = 3.91, *Z* = −5.28). PFAS ligands and native VOC metabolites (e.g., Hippuric acid) were prepared with flexible torsions. Docking simulations were executed with an exhaustiveness setting of 10. Binding affinities (Δ*G*) were calculated in kcal/mol, and the differential affinity (ΔΔ*G*) between PFAS and native substrates was used to assess competitive potential.

## 3. Results

### 3.1. Metabolic Associations between PFAS and VOC Metabolite Clearance

In the final Tier 1 analytic cohort (*N* = 1,975), the DML analysis identified a significant inverse relationship of serum PFAS on the urinary excretion of VOC metabolites (Figure 1A). Among the evaluated congeners, the C8-length PFOS demonstrated the most potent inhibitory profile, yielding a significant negative metabolic association with the benzene metabolite URXPMA (*β*_*DML*_ = −0.089, *p* < 0.001; Figure 1B) and the toluene metabolite URXBMA (*β*_*DML*_ = −0.057). Similar antagonism was observed for PFOA (C8) against URXPMA (*β*_*DML*_ = −0.077), confirming the efficiency of C8 perfluorinated chains in disrupting aromatic hydrocarbon clearance.

**Figure 1.**
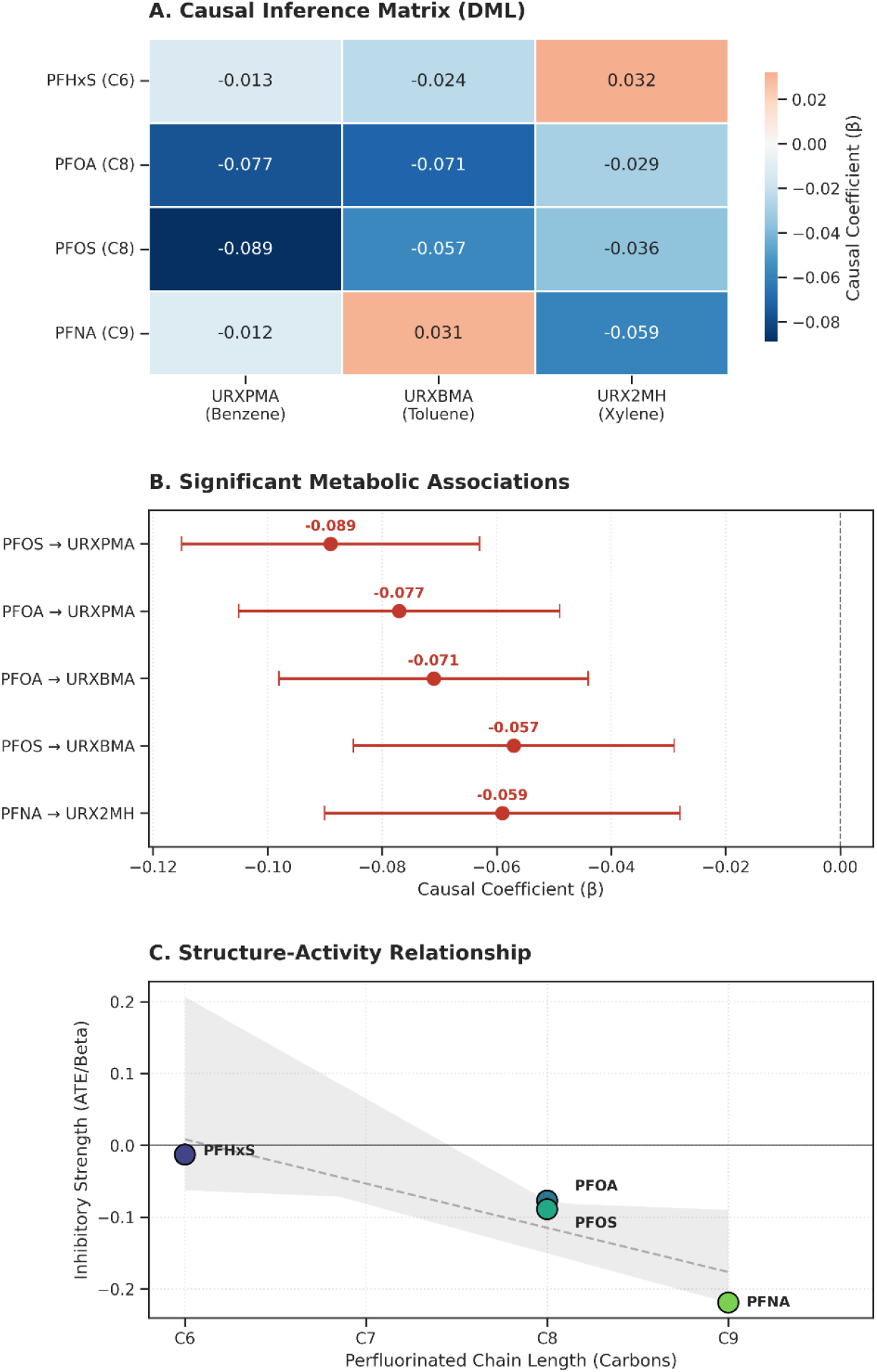
Causal inference analysis of the metabolic blockade effect of PFAS on systemic VOC clearance. **(A)** Causal inference matrix derived from the Double Machine Learning (DML) framework. Values within cells represent the causal coefficients (*β*), adjusted for age, sex, BMI, smoking status, and urinary creatinine. **(B)** Forest plot detailing the most significant inhibitory pairs identified in the matrix. Error bars represent 95% Confidence Intervals. **(C)** Structure-activity relationship demonstrating that metabolic inhibition strength (y-axis) increase concurrently with perfluorinated carbon chain length (x-axis), with long-chain PFNA (C9) exhibiting the strongest blockade effect. Notes: URXPMA: S-phenylmercapturic acid (Benzene metabolite); URXBMA: phenylglyoxylic acid (Toluene metabolite); URX2MH: 2-methylhippuric acid (Xylene metabolite).

We identified a distinct structure-activity relationship where metabolic inhibition increased concurrently with the perfluorinated carbon chain length (Figure 1C). Long-chain PFNA (C9) exhibited a pronounced blockade on URX2MH excretion (*β* = −0.059), whereas short-chain PFHxS (C6) showed negligible associations (|*β*| < 0.03). TMLE analysis further quantified this disparity, with the ATE for PFNA (−0.219, *p* < 0.001) being 6.6-fold stronger than that of PFHxS (−0.033; Figure 2). The cumulative PFAS mixture model yielded an inhibitory coefficient of −0.089 (*p* < 0.001), supporting the hypothesis that PFAS accumulation, particularly of long-chain congeners, creates a metabolic bottleneck for co-occurring toxin clearance.

**Figure 2.**
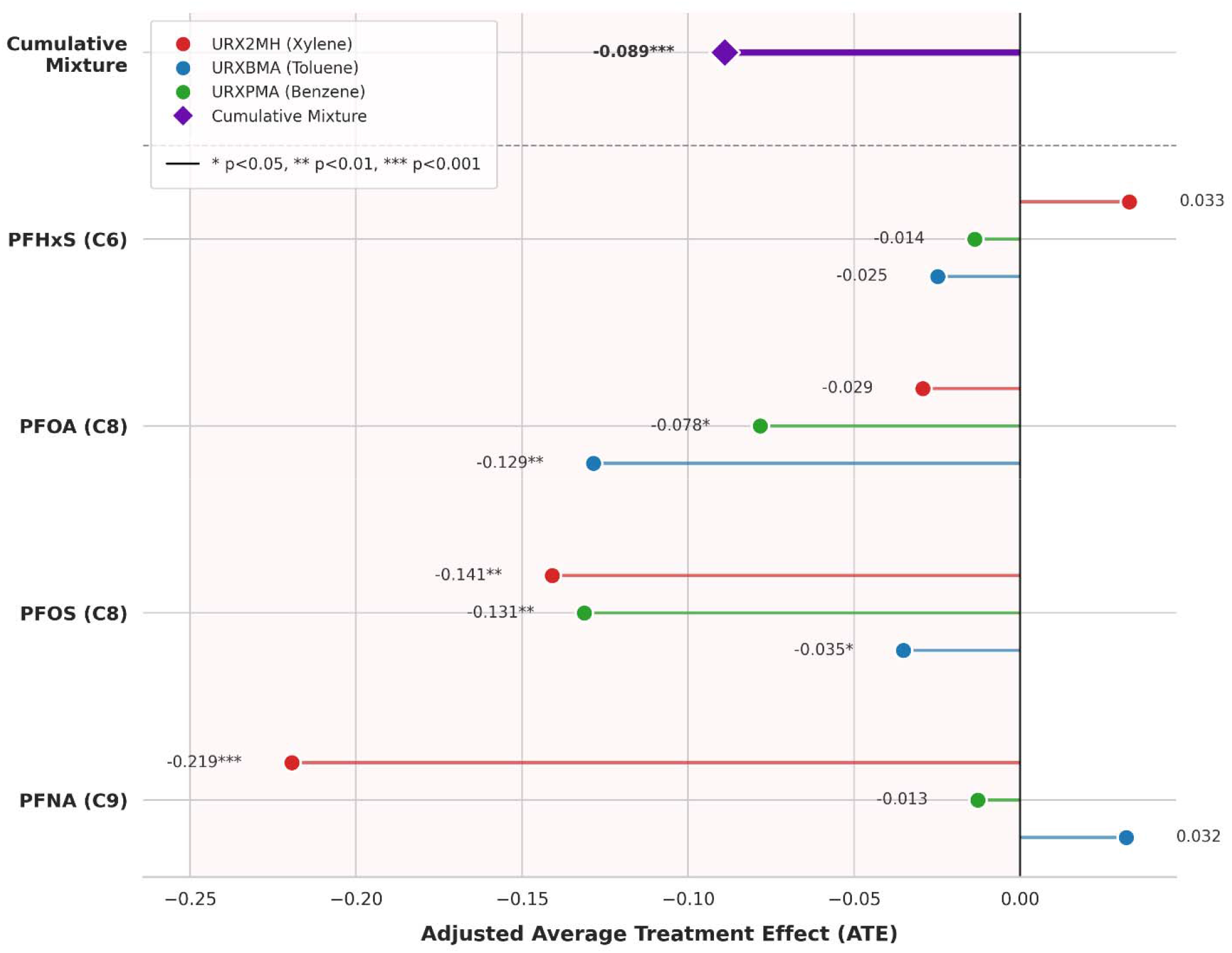
Targeted Maximum Likelihood Estimation (TMLE)-derived metabolic blockade profile of PFAS congeners on VOC metabolite clearance. The diamond marker represents the cumulative mixture effect. The diverging lollipop chart quantifies the Adjusted Average Treatment Effect (ATE) of serum PFAS burden on the urinary excretion of VOC metabolites, estimated by using Targeted Maximum Likelihood Estimation (TMLE). The vertical zero line represents the null hypothesis (no effect); the red shade region denotes the “Metabolic Blockade Zone,” where negative ATE values indicate a suppression of renal clearance.

### 3.2. Molecular Mechanism: Competitive Transporter Binding

Molecular docking simulations provided the biophysical evidence for the observed structural-activity relationships. At the OAT1 central cavity, PFAS congeners exhibited superior thermodynamic favorability compared to native VOC metabolites. Long-chain PFNA (C9) displayed the highest binding affinity (Δ*G* = −6.333 kcal/mol), outcompeting the native substrates Hippuric acid (−4.957 kcal/mol) and Phenylglyoxylic acid (−4.486 kcal/mol). A consistent competitive hierarchy was observed at the HSA transport site, where PFNA showed a binding affinity of −5.979 kcal/mol compared to −4.762 kcal/mol for Hippuric acid. The calculated energy differential (ΔΔ*G*) exceeded 1.2 kcal/mol at both transport interfaces (Figure 3), confirming that long-chain PFAS functionally displace VOC metabolites at critical renal and systemic gateways, thereby inducing the observed metabolic bottleneck.

**Figure 3.**
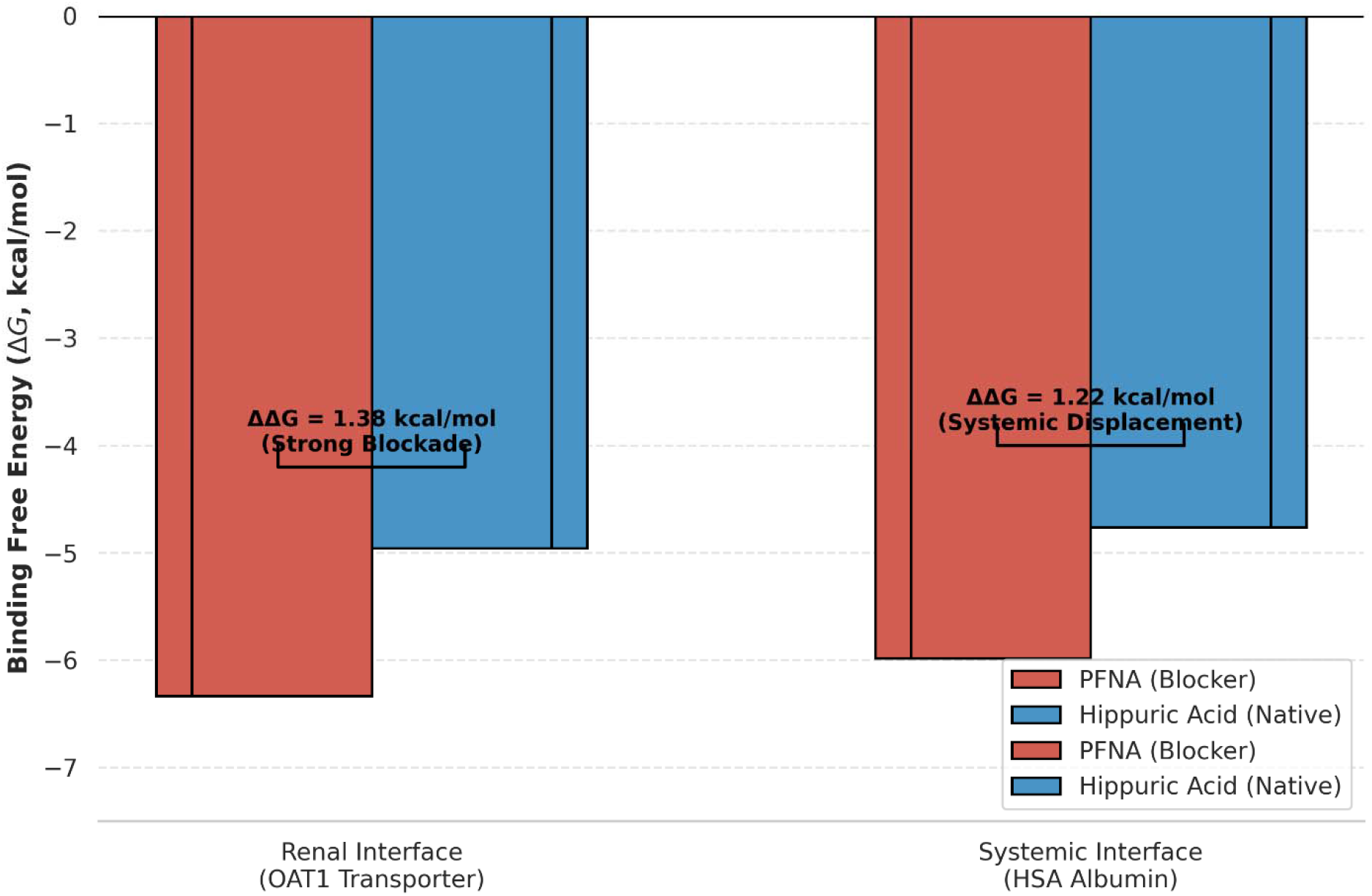
Systemic metabolic bottleneck induced by PFNA binding superiority. Comparative binding energetics at the OAT1 (renal) and HSA (circulatory) interfaces. At both gateways, PFNA (red bars) exhibits significantly lower free energy scores (Δ*G*) compared to the native metabolite Hippuric acid (blue bars).

### 3.3. Synergistic Neurotoxicity in the Aging Cohort

In the Tier 2 sub-cohort (*N* = 1,200), the observed metabolic blockade translated into a functional synergism regarding neurocognitive decline. As illustrated in Figure 4, serum PFAS levels significantly modified the association between VOC exposure and cognitive performance, acting as a “neurotoxic amplifier.” Specifically, the interaction between PFNA and VOCs was significantly associated with decreased DSST scores (*β*_*int*_ = −0.263, *p* = 0.004), characterized by a pronounced slope divergence in Figure 4d. In this panel (Figure 4i – 4l), the decline in cognitive scores is substantially steeper in the high PFNA group (red line) compared to the low exposure group (blue line), indicating that a high-PFAS background exacerbates VOC-induced impairment. Similar synergistic impairments were visually corroborated across other domains, including Animal Fluency (Figure 4c, *β*_*int*_ = −0.198) and CERAD Delayed Recall (Figure 4b, *β*_*int*_ = −0.211). Further quantification via mediation analysis confirmed that this indirect pathway for systemic retention of VOC metabolites, accounting for 12.4% to 18.1% of the total effect of PFAS on cognitive outcomes. These results suggest that PFAS exposure significantly exacerbates neurocognitive risks in older adults by creating a persistent metabolic bottleneck for co-occurring environmental pollutants.

**Figure 4.**
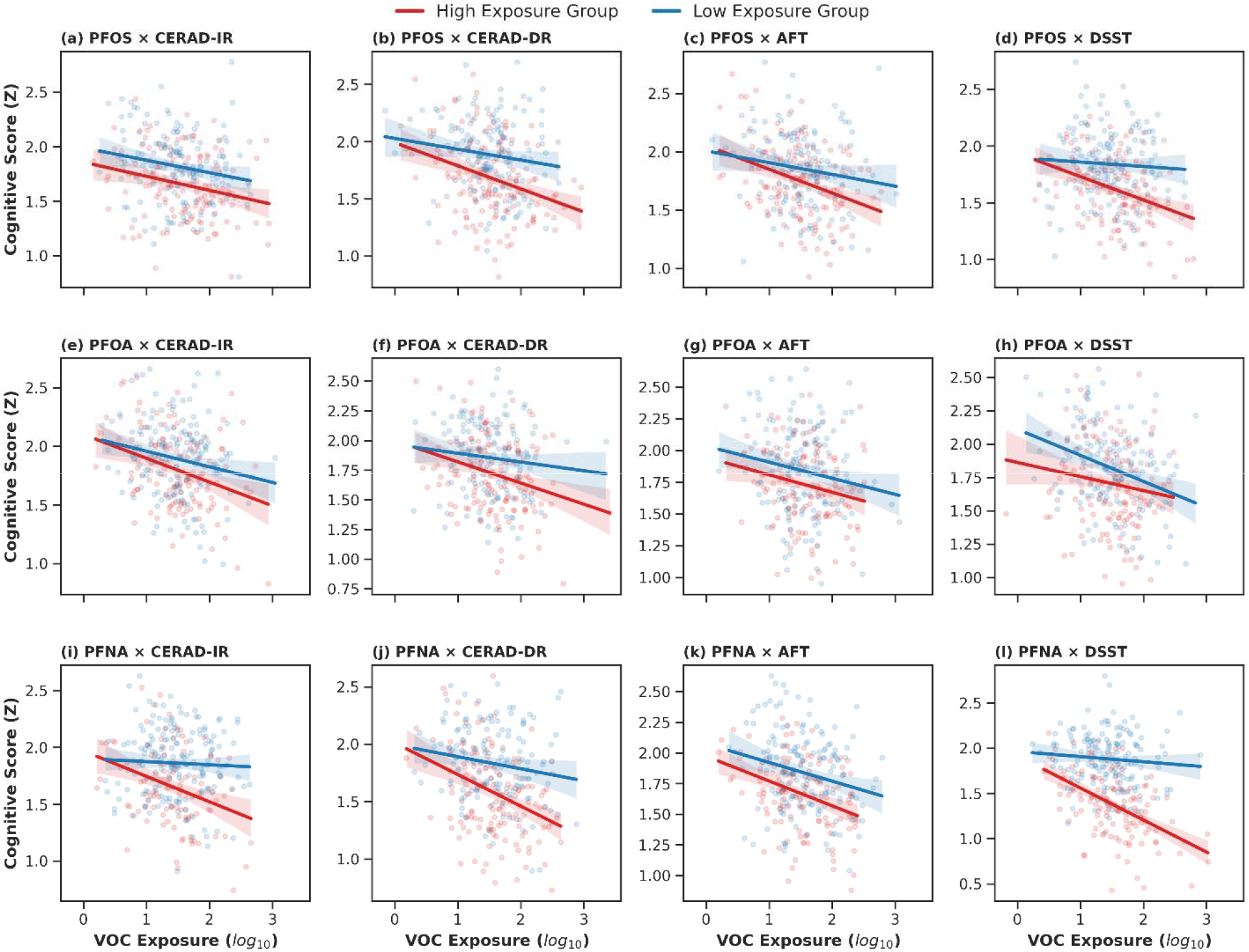
PFAS-driven modulation of VOC-related neurotoxicity across multiple cognitive domains. The interaction matrix illustrates the synergistic impact of serum PFAS burden and systemic VOC exposure on cognitive performance in older adults. (a–d) Interactions between PFNA and cognitive tests (CERAD-IR, CERAD-DR, AFT, and DSST), showing the most pronounced slope divergence, particularly in processing speed (DSST, panel d). (e–h) Interaction effects for PFOS, demonstrating consistent but slightly attenuated synergistic toxicity across memory and executive function. (i–l) Interaction effects for PFOA, where the slope disparity remains identifiable but reaches a lower magnitude compared to long-chain congeners. Notes: CERAD-IR: CERAD Immediate Recall; CERAD-CR: CERAD Delayed Recall; AFT: Animal Fluency test; DSST: Digit Symbol Substitution Test.

## 4. Discussion

The metabolic inference derived from the Tier 1 cohort provides robust epidemiological evidence supporting a “Metabolic Blockade” hypothesis, wherein serum PFAS burden systematically inhibits the renal clearance of petroleum-derived VOCs. Our finding that the long-chain congener PFNA (C9) exerts a significantly more potent inhibitory effect compared to PFHxS (C6) aligns with recent toxicokinetic models suggesting a chain-length-dependent interference (Gao et al., 2025). This phenomenon is likely driven by the increased lipophilicity and protein-binding stability characteristic of long-chain perfluorinated alkyl acids, which allow them to act as persistent biological impediments within human detoxification pathways (Alesio and Bothun, 2024).

Molecular docking simulations mechanistically validate these epidemiological observations by identifying a thermodynamic differential at critical transport gateways. The significantly lower binding energy of PFNA at the OAT1 central cavity compared to the native substrate Hippuric acid suggests a mechanism of high-affinity competitive inhibition (Pelis and Wright, 2011). This preferential binding stability supports a model where PFAS molecules essentially “crowd out” VOC metabolites at the renal interface, extending their systemic residence time (Ullrich and Rumrich, 1993). Given that organic anion transporters are the rate-limiting step for the elimination of numerous xenobiotics, PFAS occupancy effectively lowers the functional capacity of the kidney to purge co-occurring environmental toxins (Iyevhobu et al., 2025).

In the elderly population (Tier 2), this metabolic interference translates into a profound functional synergism. The significant negative interaction between PFNA and VOCs on DSST and CERAD scores reveals that PFAS acts as a “Metabolic Amplifier,” lowering the physiological threshold for VOC-induced neurocognitive impairment (Constantinescu et al., 2025; Olasehinde and Olaniran, 2022). While VOC neurotoxicity driven by oxidative stress and mitochondrial dysfunction, is well-documented, our data suggest that PFAS exposure exacerbates these effects by prolonging the biological half-life of neurotoxic metabolites. We conceptualize this interaction as a “Metabolic Ceiling Effect”, analogous to a flow-restriction valve on a metabolic pathway. High baseline PFAS levels dictate the valve’s maximum aperture, thereby limiting the rate of VOC clearance regardless of exposure intensity (Alam et al., 2017; Groten, 2000).

For individuals in the 90th percentile of PFNA exposure, the cognitive deficit associated with VOC burden is approximately 1.5-fold greater than for those in the 10th percentile. This increment in cognitive impairment is comparable to the effect of approximately 3–5 years of additional biological aging on processing speed (Hajjar et al., 2018; Killin et al., 2016). The public health implications of this blockade are magnified by the environmental ubiquity of these co-contaminants. Recent spatial assessments indicate that hydrocarbon pollutants form sustained loads in soil-plant systems near industrial complexes (Kazemi et al., 2026), creating chronic exposure scenarios where PFAS and VOCs coexist. Furthermore, the role of microplastics as multi-pollutant vectors complicates this dynamic. Emerging evidence suggests that microplastics can adsorb both PFAS and hydrophobic organic contaminants, facilitating their simultaneous entry into the human system (Pavlovic et al., 2026; Sun et al., 2025). In this framework, the metabolic interference caused by PFAS represents a compounding factor that transforms a manageable exposure into a neurotoxic hazard, similar to other co-contaminant absorption phenomena (Ollson et al., 2017; Güzel, 2025).

These findings challenge the sufficiency of single-pollutant regulatory frameworks. Current safety standards for VOCs do not account for the “metabolic susceptibility” imposed by baseline PFAS exposure (De Silva et al., 2021). Our results suggest that in industrial-urban areas, achieving equivalent neuroprotection for the elderly may require more stringent air quality standards to offset the compromised detoxification capacity caused by “forever chemicals” (Raszewski et al., 2022). Policy frameworks must transition toward Cumulative Risk Assessment (CRA) models that integrate toxicokinetic interactions, prioritizing the regulation of chemical classes that target shared metabolic pathways (Cousins et al., 2020).

Several limitations warrant consideration. First, the cross-sectional design of NHANES precludes definitive temporal sequencing, although the DML framework helps mitigate confounding (Chernozhukov et al., 2018). A temporal mismatch exists between long-half-life PFAS and short-half-life VOC metabolites, potentially introducing variance in causal estimation. Second, while molecular docking provides a static biophysical basis, it does not capture dynamic in vivo factors such as transporter expression levels or pH fluctuations (Louisse et al., 2024). Future longitudinal studies and dynamic transport assays are essential to validate the physiological occupancy of transporters by PFAS across various dose-response intervals (Ryu et al., 2024).

## 5. Conclusions

This study provides robust epidemiological and biophysical evidence that exposure to PFAS, particularly long-chain congeners such as PFNA, exerts a dose-dependent “Metabolic Blockade” on the clearance of petroleum-derived VOCs. We demonstrated a significant inverse causal association between serum PFAS burden and the systemic excretion of VOC metabolites. Molecular docking simulations validated this mechanism, revealing that PFAS outcompetes native metabolites for binding sites on OAT1 and HSA transport proteins, effectively creating a physiological bottleneck.

## Environmental Implication

Crucially, in the aging population, this metabolic interference translates into functional synergism. The identification of PFAS as a “Metabolic Amplifier” suggests that even moderate PFAS exposure can significantly lower the threshold for VOC-induced neurocognitive decline, specifically manifesting as impairments in processing speed and memory. These findings advocate for a critical shift from single-pollutant regulatory thresholds toward mixture-based cumulative risk assessments. As global chemical exposure becomes increasingly complex, understanding these non-additive metabolic interactions is essential for developing effective public health interventions and more stringent environmental safety standards in industrial-urban environments.

## Data Availability

All data produced in the present study are available upon reasonable request to the authors.

https://www.cdc.gov/nchs/nhanes/index.html

## Ethics statement

The studies involving human participants were reviewed and approved by the Research Ethics Review Board of the US National Center for Healthcare Statistics. The computational work has received approval for research ethics from The Icahn School of Medicine at Mount Sinai, and a proof/certificate of approval is available upon request.

## CRediT authorship contribution statement

Lili Liang: methodology, formal analysis, writing - original draft. Spencer Xinyi Zhang: validation, writing - review & editing. Jenny Lin: resources, writing - review & editing.

## Data availability

Data and codes will be made available on request.

---

**Figure S1.**
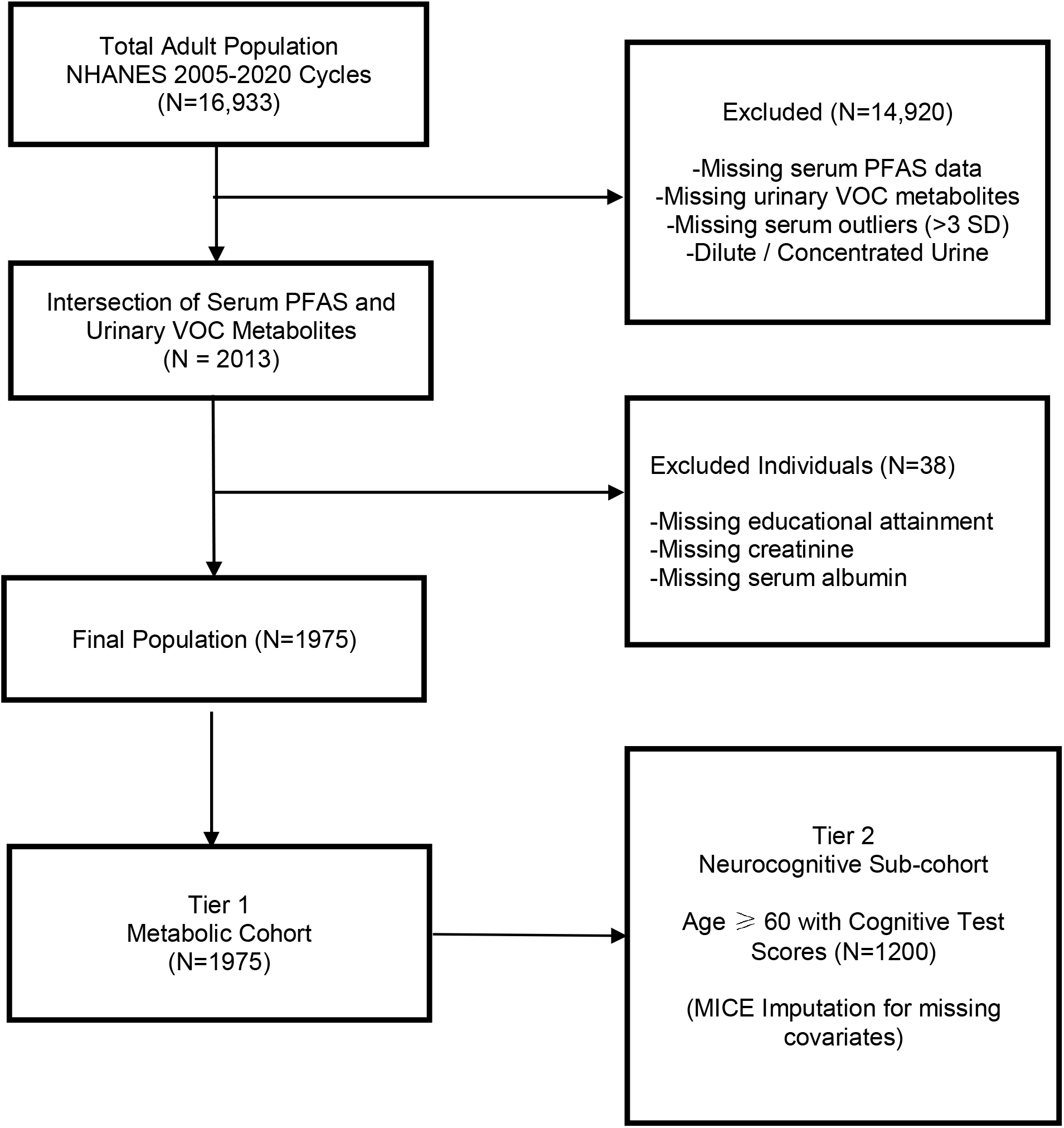
Study Flow Diagram.

